# Time-Restricted Eating and Dietary Intake Shape 24-Hour Glycemic Patterns in Type 2 Diabetes: A Functional Data Analysis Approach

**DOI:** 10.1101/2025.07.26.25332240

**Authors:** Wenxin Xu, Collin Sakal, Wei Zhang, Tong Chen, Congrong Wang, Qinpei Zhao, Xinyue Li

## Abstract

**Background:** Time-restricted eating (TRE) shows promise for metabolic health, but its effectiveness in glucose management among individuals with type 2 diabetes, and its role in modulating glucose responses to dietary intake remain poorly understood.

**Objective:** We aimed to identify temporal associations of TRE and dietary intake with 24-hour glucose dynamics. We further examined the interactions between eating windows and carbohydrate intake.

**Methods:** Clinical information, dietary records, and continuous glucose monitoring data from 90 Chinese adults with type 2 diabetes were analyzed. Two digital biomarkers were developed to characterize duration and regularity of eating patterns: a binary indicator for eating windows <10 hours (TRE10) and a continuous measure of deviation from an individual’s median eating window (TWD). Linear mixed-effects models and functional data analysis were used to examine independent and temporal associations of glucose levels with TRE and dietary intakes.

**Results:** TRE10 was associated with reduced mean amplitude of glycemic excursions (MAGE) (β = −7.15, P = 0.031) and glucose standard deviation (SD) (β = −2.16, P = 0.035), with the strongest associations around 09:00. TWD was positively associated with glycemic coefficient of variation (CV) (β = 0.36, P = 0.050) and higher glucose levels between 07:00 and 08:00. Carbohydrate intake was significantly associated with time in range (TIR) and glycemic variability, with notable glucose changes at 10:00 and 21:00. Dietary vitamin D intake was linked to reduced glycemic area under the curve (AUC) (β = −0.29, P = 0.040), with pronounced effects at 12:00 and 20:00. Additionally, eating windows < 10 hours attenuated carbohydrate-induced glucose spikes in the morning but amplified glucose responses in the early afternoon.

**Conclusions:** In adults with type 2 diabetes, eating windows <10 hours and consistent eating windows improved glycemic control, with distinct time-of-day effects. These findings support integrating timing-based nutritional strategies into personalized diabetes management.

## Introduction

Type 2 diabetes presents a major global health challenge, with its prevalence projected to reach 783.2 million by 2045 (1). In China, culturally specific dietary patterns high in carbohydrates contribute to the country’s status as having the largest diabetes population worldwide (2). Dietary interventions are a fundamental component of first-line therapeutic approaches for type 2 diabetes management (3). Although conventional calorie-restricted diets have demonstrated efficacy in improving glycemic levels, their long-term adherence remains a major challenge due to the complexities of calorie tracking (4). Time-restricted eating (TRE) has emerged as a promising alternative by focusing on eating window limitation rather than calorie counting. Studies conducted among participants without diabetes suggest that TRE can enhance glucose regulation (5, 6). Additionally, modern irregular eating patterns and unrestricted food availability often disrupt circadian rhythms, contributing to metabolic health issues (7). TRE has shown potential in restoring circadian alignment (8). However, the effectiveness of TRE in individuals with type 2 diabetes remains poorly understood, and to our knowledge, no study has examined the interactions between TRE and dietary intake on glucose levels in this population.

Effective management of type 2 diabetes requires strategies that address not only chronic hyperglycemia but also glycemic variability, which have been linked to an increased risk of cardiovascular complications (9). Despite its importance, glycemic variability is often overlooked in practice, where Hemoglobin A1c (HbA1c) serves as the primary biomarker. While HbA1c is a reliable indicator of average glucose levels, it fails to capture short-term glucose fluctuations and acute excursions (10). Continuous glucose monitoring (CGM) has demonstrated its effectiveness in diabetes management (11, 12), offering a powerful tool to overcome the limitations of traditional glycemic biomarkers. However, conventional CGM-derived metrics, such as in-range and variability indices, reduce glucose data into summary statistics that fail to fully capture the complexity of temporal glycemic dynamics (13). Advanced analytical approaches, such as functional data analysis (FDA), offer a novel framework for modelling glucose data as continuous trajectories over time, enabling a detailed understanding of glycemic patterns.

In this study, we investigated the association of time-restricted eating and dietary intake with glycemic profiles among Chinese adults with type 2 diabetes under free-living conditions. Leveraging multi-modal data, including clinical information, food records, and CGM data, we applied advanced statistical methods to analyze temporal associations of time-restricted eating and nutritional content with glucose dynamics. We further examined the potential modulating effects of eating windows on glycemic responses to carbohydrate intake. This work provides new insights into how eating patterns and nutrient composition of are associated with blood glucose levels, highlighting the potential for personalized TRE-based and dietary strategies to improve glycemic management.

## Methods

### Study design and population

Data for this study were obtained from the ShanghaiT2DM Dataset, a publicly available dataset collected in Shanghai, China (14). The dataset includes 100 participants diagnosed with type 2 diabetes, all aged 18 years or older. Participants were recruited from the Diabetes Data Registry and Individualized Lifestyle Intervention (DiaDRIL) program at Shanghai Fourth People’s Hospital (June 2021 to November 2021) and Shanghai East Hospital (September 2019 to March 2021). All participants wore FreeStyle Libre flash continuous glucose monitors (Abbott Diabetes Care, Witney, UK) to measure interstitial glucose levels over recording periods ranging from 3 to 14 days, with measurements taken at 15-minute intervals. A total of 112,475 CGM measurements were collected. Self-reported meal intake data was recorded under free-living conditions, including meal timings and weighted food records. In addition, the ShanghaiT2DM dataset includes demographic information, anthropometric measurements, and clinical test results obtained during hospital visits via questionnaires and medical records. This study utilized anonymized, publicly available data from the ShanghaiT2DM dataset, which was ethically approved by relevant ethics committees and obtained with informed consent.

For this study, data from multiple modalities were analyzed, including CGM sensor data, demographic information, and clinically documented medical examinations. Missing CGM records of no more than 2 hours were imputed using linear interpolation. Data cleaning was performed at the day level to align the modalities. Days were excluded if they met any of the following criteria: fewer than 96 CGM recordings after imputation, missing covariates, or incomplete meal recordings.

### Nutritional composition and TRE digital biomarkers

A total of 3,061 self-reported, non-empty meal records were collected in the ShanghaiT2DM dataset. Each food ingredient was mapped to a standardized nutrient database to derive its nutritional content, including carbohydrate, protein, lipid, fiber, and vitamin D. Nutritional values for meals were calculated by summing the nutrient contents of all ingredients and adjusting proportionally based on reported food quantities. The nutritional information was obtained from FoodData Central from the U.S. Department of Agriculture (15), supplemented by two culturally relevant tools: Boohee App and the Centre for Food Safety, Hong Kong (16).

Meal timings were self-reported and aligned with CGM recordings. Food records reported within 30 minutes were merged. Meals with extreme nutritional content were excluded based on the following thresholds: >1,500 kcal, >1,000 grams of carbohydrates, >1,000 grams of protein, >100 grams of lipid, >100 grams of dietary fibers, or >100 micrograms of dietary vitamin D. Processed meal-level data were aggregated at the daily level to compute total nutrient intake per participant per day. To ensure consistency with the three-meal dietary structure typical in Chinese culture and to avoid missing records, only days with at least three recorded meals were included in the analysis.

Two digital biomarkers were introduced to characterize time-restricted eating: a binary indicator for eating windows <10 hours (TRE10) and a continuous measure of deviation from an individual’s median eating window (TWD). The daily eating window was calculated as the time elapsed between the first and last meal of the day. Consistent with two previous clinical trials in individuals with type 2 diabetes, a 10-hour threshold was used to define TRE10 (17, 18). Days were classified as having a “short” eating window (<10 hours) or a “long” eating window (≥10 hours). TWD (Time Window Difference) was computed in two steps. First, for each participant, the median eating window across all study days was calculated as a personalized reference. Second, for each day, TWD was calculated as the difference between the observed daily eating window and the participant’s median eating window. The median eating window was chosen over the mean due to its robustness against outliers.

### Standard CGM-derived summary statistics analysis

Day-based summary statistics were derived from the CGM data to capture both in-range metrics and glucose variability features. Specifically, the glycemic summary statistics included the percentage of time glucose levels were between 70 and 180 mg/dL (TIR), the percentage of time glucose levels exceeded 180 mg/dL (TAR), the percentage of time glucose levels were below 70 mg/dL (TBR), the hourly average area under the glucose curve (AUC), the coefficient of variation (CV), the mean amplitude of glycemic excursions (MAGE), and the standard deviation (SD). Linear mixed-effects models with random intercepts were employed to assess the independent associations of time-restricted eating and nutritional content with each glycemic summary statistic. All models were adjusted for potential confounders, including age, sex, diabetes duration, body mass index, smoking status, and alcohol consumption.

### Functional data analysis

Given the continuous nature of CGM data, consecutive measurements are naturally more dependent than independent of each other. To fully leverage the rich information in CGM recordings, we applied FDA to capture the detailed temporal effects of TRE and nutritional intake on CGM trajectories. Specifically, we employed Fast Univariate Inference to conduct epoch-by-epoch function-on-scalar regression (19), modelling CGM data as functional outcomes and exposures and covariates as scalar variables. The analysis proceeded through epoch-wise linear mixed-effect models, with random intercepts that accounting both within- and between-subject variability, explicitly incorporating the multilevel structure of CGM data. CGM recordings, collected at 15-minute intervals, resulted in 96 epochs per day. As recommended by Crainiceanu et al. (20) coefficient trajectories were smoothed using locally weighted scatterplot smoothing (LOWESS) with a span parameter of 1/24 for one-hour interval. Confidence intervals were derived using bootstrapping with 1,000 iterations.

We first examined the temporal associations between nutritional intake and CGM levels, adjusting for age, sex, diabetes duration, body mass index, smoking status, and alcohol consumption. Next, we investigated the temporal effects of two TRE digital biomarkers, TRE10 and TWD, while adjusting for nutritional intake. To explore whether eating window duration modulates the glycemic response to carbohydrate intake, we introduced an interaction term between daily carbohydrate consumption and TRE10 in the fully adjusted model.

### Sensitivity analysis

Sensitivity analyses were performed to assess the robustness of our findings. We conducted subgroup analyses stratified by sex (males and females) and age (older group ≥63 years; younger group <63 years). The age cutoff of 63 years was chosen based on the median age of the study cohort. The associations between eating windows and nutritional components with CGM levels were evaluated within each subgroup using the same statistical methods as in the main analysis, except for females, where smoking and alcohol drinking were not adjusted, as all females were non-smokers and non-drinkers. The consistency of the direction and significance of the associations across subgroups was compared to the main analysis results.

## Results

### Baseline characteristics

In this study, we included 90 Chinese adults from the ShanghaiT2DM Dataset (46% female; mean age 60.2 ± 13.9 years; HbA1c 76.0 ± 27.1 mmol/mol; diabetes duration 8.9 ± 8.6 years; **Table 1**). Over an average continuous glucose monitoring period of 10.0 ± 3.3 days, participants contributed 65,280 glucose measurements across 680 participant-days, with glucose values recorded at 15-minute intervals. Complete baseline clinical and demographic characteristics alongside CGM adherence are detailed in **Table 1** at participant level, and an inclusion flowchart is presented in **Supplementary Figure 1**. Mean daily caloric intake was 1072.3 ± 469.8 kcal, with dietary patterns characterized by macronutrient composition (carbohydrates, protein, lipids, and fiber) and dietary vitamin D intake.

**Table 1.** Baseline characteristics of participants.

| Characteristic | Mean (SD/%) |
| --- | --- |
| Age (years) | 60.22 (13.91) |
| Sex |  |
| Female | 41 (45.56%) |
| Male | 49 (54.44%) |
| Duration of diabetes (years) | 8.87 (8.56) |
| BMI (kg/m<sup>2</sup>) | 24.06 (3.13) |
| Smoking |  |
| Non-smoker | 76 (84.44%) |
| Smoker | 14 (15.56%) |
| Alcohol |  |
| Drinker | 9 (10.00%) |
| Non-drinker | 81 (90.00%) |
| FPG (mg/dL) | 169.98 (66.59) |
| PPG (mg/dL) | 268.94 (100.82) |
| HbA1c (mmol/mol) | 75.99 (27.09) |
| CGM Recording period (days) | 9.98 (3.30) |

BMI, body mass index; CGM, continuous glucose monitoring; FPG, fasting plasma glucose; HbA1c, Hemoglobin A1c; PPG, 2-hour postprandial plasma glucose.

### Digital biomarkers for time-restricted eating

Among participants with valid data, the mean ± SD eating window was 10.7 ± 1.5 hours with a range of 7.5–19 hours. The mean time of the first recorded meal was 07:23, and the mean time of the last recorded meal was 18:07. To evaluate the potential impact of TRE on glucose profiles, we introduced two daily digital biomarkers: (i) TRE10 (Time-Restricted Eating <10h), a binary indicator representing eating windows <10 hours, and (ii) TWD (Time Window Deviation), a continuous variable that quantifies the difference between the observed daily eating window and the participant’s median eating window (see “Methods”). Complete day-level characteristics, including dietary factors and CGM-derived glucose measures, are presented in **Table 2**.

**Table 2.**
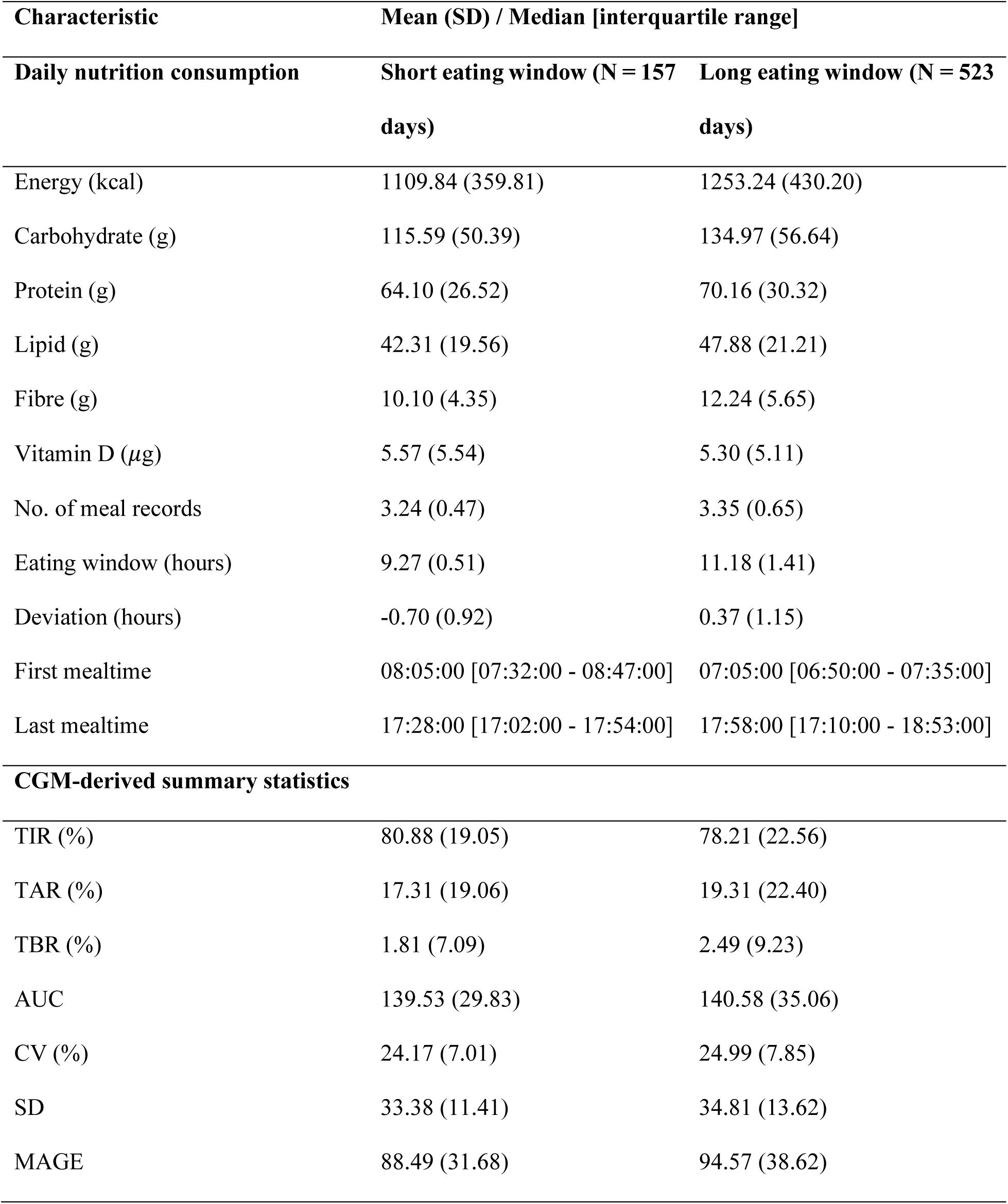
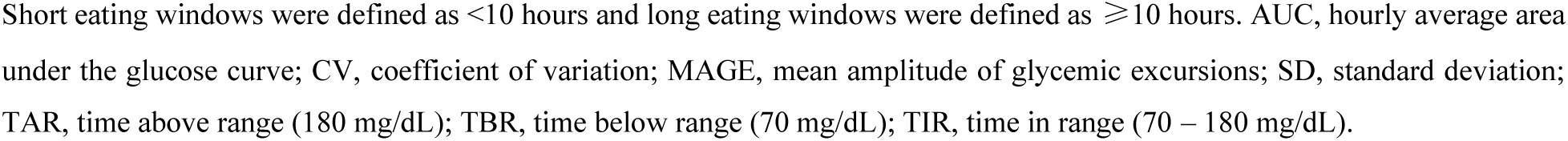
Day-level nutrition content and CGM-derived statistics summary categorized by eating windows duration.

### Independent associations of TRE and dietary intake with CGM summary statistics

The independent associations of TRE and dietary intake with glycemic control were analyzed using adjusted linear mixed-effects models, accounting for both between- and within-subject variability. The analysis included a range of CGM-derived summary statistics, comprising in-range as well as variability metrics. Having an eating window ≤ 10h (TRE10) was significantly associated with lower glycemic variability, measured by mean amplitude of glycemic excursions (MAGE) and glucose standard deviation (SD) (MAGE, β = −7.14, P = 0.031; SD, β = −2.16, P = 0.035). In contrast, the deviation in eating windows (TWD) showed a positive association with glucose variability as measured by coefficient of variation (CV) of glucose levels (β = 0.36, P = 0.050). Carbohydrate consumption was significantly associated with the percentage of time glucose levels between 70 and 180 mg/dL (TIR, β = 0.31, P = 0.029) and percentage of time glucose levels exceeding 180 mg/dL (TAR, β = 0.28, P = 0.039). Additionally, it was associated with variability measures, including the coefficient of variation (CV, β = 0.16, P = 0.005), the mean amplitude of glycemic excursions (MAGE, β = 0.71, P = 0.018), and the standard deviation (SD, β = 0.27, P = 0.005). Dietary vitamin D intake was negatively associated with hourly average area under the glycemic curve (AUC, β = −0.29, P = 0.040). Daily intake of dietary protein, lipid, and fiber was not significantly associated with any CGM-derived summary statistics (**Figure 1**, **Supplementary Table 1**).

**Figure 1.**
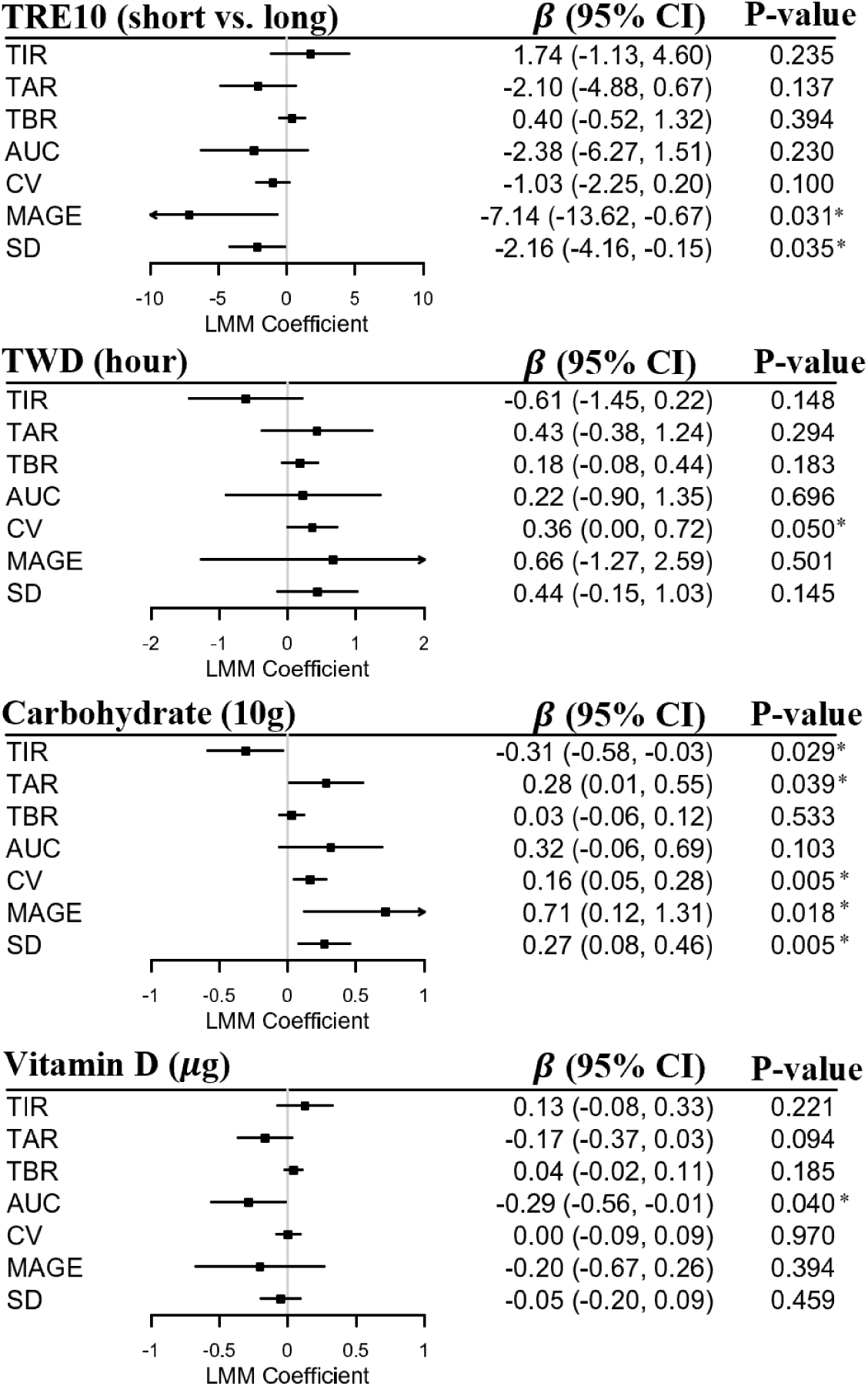
Independent associations of time-restricted eating and nutritional contents with CGM-derived summary statistics. The black squares and horizontal lines represent estimated coefficients and 95% CIs respectively in linear mixed-effect models. TRE10 defined eating windows into “short” (<10 hours) and “long” (≥10 hours), with the “long” eating window group serving as the reference. TWD defined as the time difference between the observed daily eating window and the participant’s median eating window. Asterisks identify statistical significance (P < 0.05). Daily carbohydrate and dietary vitamin D intakes were obtained from dietary records. The models were adjusted for age, sex, diabetes duration, BMI, alcohol, and smoking. AUC, hourly average area under the glucose curve; CV, coefficient of variation; MAGE, mean amplitude of glycemic excursions; SD, standard deviation; TAR, time above range (180 mg/dL); TBR, time below range (70 mg/dL); TIR, time in range (70 – 180 mg/dL). Asterisks identify statistical significance (P < 0.05).

### Temporal associations of TRE and dietary intake with functional CGM data

To investigate the dynamic temporal effects of TRE and dietary intake on glucose levels, function-on-scalar regression analysis was performed (**Figure 2**). TRE10 was associated with decreases in CGM levels, with the most substantial reductions observed around 09:00 and smaller decreases between 16:00 and 17:00. TWD showed more modest effects, with the most evident positive associations occurring between 07:00 and 08:00. The morning effects of both TRE10 and TWD were more pronounced, as reflected by narrower confidence intervals. Daily carbohydrate consumption was primarily associated with breakfast postprandial hyperglycemia, with peak effects observed at 10:00 and 21:00. Lipid intake was associated with elevated glucose levels in the afternoon, primarily between 17:00 and 19:00. Dietary fiber was negatively associated with glucose levels around 12:30. Dietary vitamin D showed negative associations with glucose levels at 12:00 and 20:00. Minimal effects were observed for all nutritional variables during the nocturnal fasting period (00:00–06:00), except for protein, which demonstrated a weak positive association with glucose levels around 03:00. The interaction between TRE10 and carbohydrate intake, the primary nutritional determinant of CGM values, was also examined. TRE10 weakly attenuated carbohydrate-induced glycemic responses around 11:00. However, despite TRE10’s overall protective effects on glycemic profiles, the interaction effect showed positive associations during the afternoon period (14:00–15:00), indicating an amplified glucose response to carbohydrate intake during this time.

**Figure 2.**
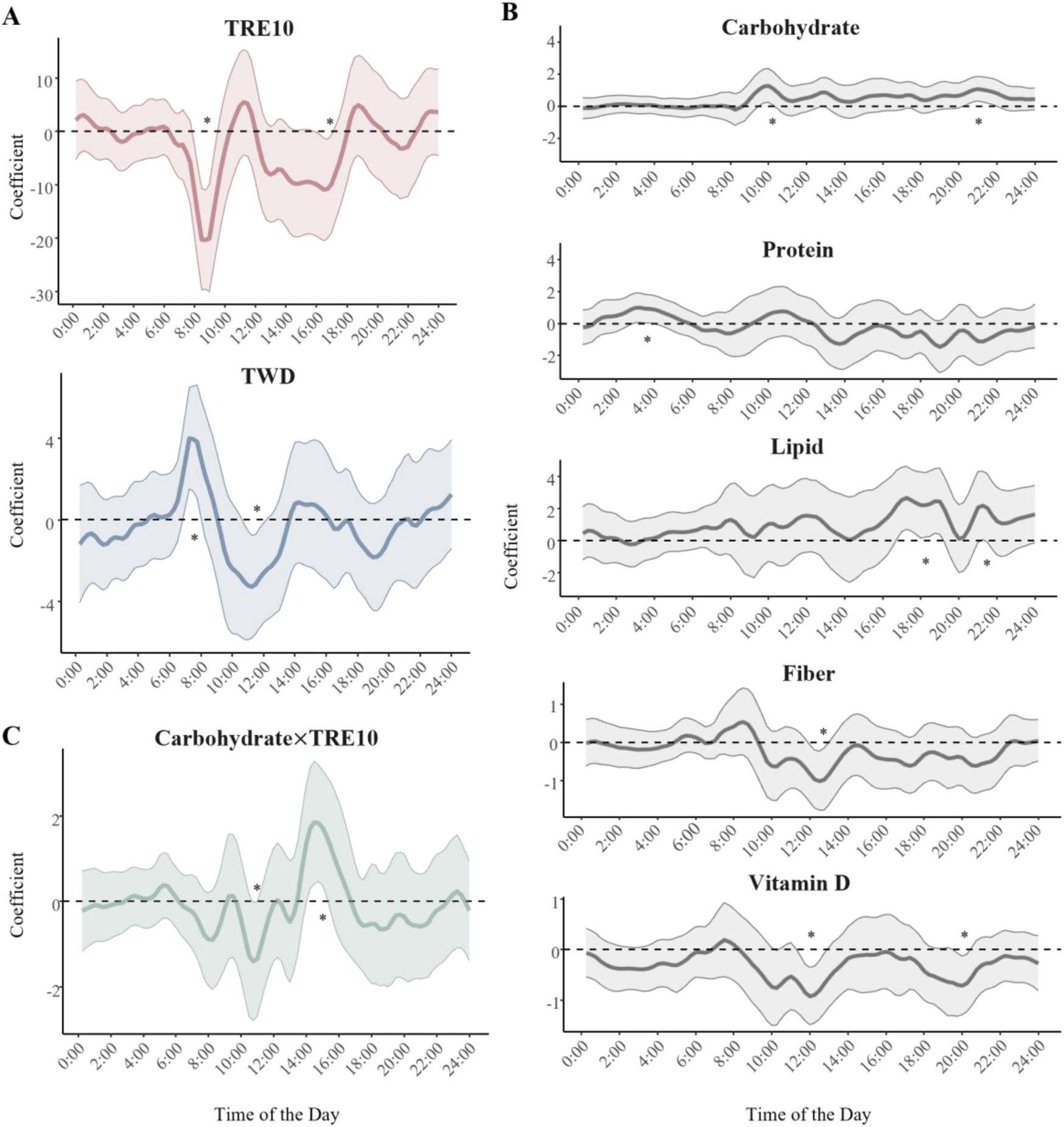
Temporal associations of time-restricted eating and nutritional contents with CGM levels. The solid curves and shaded area represent estimated coefficients and 95% CIs respectively in function-on-scalar regression and bootstrapping. Dashed horizontal lines represent coefficients equal to zero. Asterisks identify statistical significance using 95% CIs. A: TRE10 defined eating windows into “short” (<10 hours) and “long” (≥10 hours), with the “long” eating window group serving as the reference. TWD defined as the time difference between the observed daily eating window and the participant’s median eating window. B: Daily dietary nutritional contents. C: Interaction between TRE10 and daily carbohydrate intake. All models were adjusted for age, sex, diabetes duration, alcohol and smoking.

### Sensitivity analysis

Sensitivity analyses confirmed the robustness of our findings. Subgroup analyses by sex revealed that eating windows within 10 hours were significantly associated with increased glycemic variability in males, with a similar, though non-significant, trend observed in females (**Supplementary Tables 2, 3**). Deviation from the median eating window showed moderate associations with glycemic variability in males, while no evident associations were observed in females. Carbohydrate intake consistently correlated with deteriorated in-range measures and increased glycemic variability across both sexes, while dietary vitamin D intake demonstrated associations in the same direction as the main analysis. Other nutritional components exhibited slightly different effects between males and females, likely due to reduced sample sizes. Further subgroup analysis by age (older group ≥63 years; younger group <63 years) produced consistent results with the main analysis. Although associations between eating windows and glucose levels were not statistically significant in the older group, the direction of effects remained consistent. Carbohydrate intake and dietary vitamin D displayed similar associations in both age subgroups (**Supplementary Tables 4, 5**).

## Discussion

In this observational cohort study, we introduced two digital biomarkers, TRE10 and TWD, to quantify eating windows and their variability, providing a novel framework to evaluate the potential effectiveness of time-restricted eating among a Chinese cohort with type 2 diabetes under free-living conditions. The findings suggest that these metrics could provide valuable insights into glycemic variability control. Through the integration of CGM data and FDA, the temporal effects of eating patterns and nutritional intake on glycemic levels were investigated, identifying distinct glycemic patterns at different times of the day. Additionally, our analysis revealed a complex interaction between carbohydrate intake and eating windows, suggesting that both the timing and composition of meals play critical roles in glycemic control. While the study design does not allow for causal conclusions, these findings underscore the potential clinical and public health significance of integrating eating time windows and nutritional content into strategies for diabetes management.

Our study extends the application of TRE to individuals with type 2 diabetes, and introduces two novel digital biomarkers, TRE10 and TWD, to quantify eating patterns and their variability in free-living conditions. Earlier studies demonstrated that TRE could improve metabolic biomarkers in individuals without diabetes (5, 6). However, the relevance of these findings to type 2 diabetes has remained limited due to the distinct pathophysiology of diabetes and the added complexity of glycemic control in this population (21). In alignment with clinical trials conducted in participants with type 2 diabetes (17, 18), we used a 10-hour cutoff to define the digital biomarker TRE10. Building on evidence that variability in meal timing adversely affects glycemic regulation (22), we introduced TWD as a novel measure to quantify the consistency of eating patterns. To further account for individual differences in chrono-nutrition influenced by an individual’s chronotype (23), we calculated TWD using personalized median eating windows. By advancing digital biomarkers to assess both eating window duration and regularity, our study contributes to the growing field of chrono-nutrition.

We found significant associations between TRE10 and measures of glycemic variability and provided the first evidence that the glucose-lowering effect of a short eating window was most pronounced during the morning hours. While the international consensus has emphasized the utility of CGM devices for assessing glycemic outcomes (24), few clinical trials have incorporated CGM-derived metrics (18, 25, 26). Among these, one existing study notably reported a reduction in median absolute deviation with a 9-hour eating window (25), which aligns with our findings that shorter eating windows reduce glycemic variability. For in-range CGM metrics, our results are consistent with the prior study that reported no significant increase in TIR after a 6-month intervention (26). In contrast, two other trials observed significant increases in TIR (18, 25), potentially due to shorter intervention durations of 3 weeks (18) and 4 weeks (25), compared to the longer 6-month intervention (26) and the habitual observation period in our study. These findings suggest that shorter interventions may capture initial physiological adaptations to TRE, while longer durations are necessary to evaluate its sustained effects on glycemic control in free-living conditions. This distinction is critical for designing future studies and translating TRE into clinical practice. Furthermore, given that β-cell responsiveness exhibits a circadian peak in the morning (27), it is physiologically plausible that the observed benefits of TRE10 are most evident during this time of day. Overall, our findings emphasize the importance of both the duration and regularity of eating windows in shaping glycemic outcomes and suggest that morning-focused TRE strategies may improve glucose regulation in individuals with type 2 diabetes.

Greater daily irregularity in habitual eating windows, measured by TWD, was significantly associated with increased glycemic variability, with FDA revealing an adverse impact in early mornings. This finding supports and extends existing evidence on the negative effects of irregular eating patterns and circadian misalignment on metabolic health (28). Recent study has also linked irregular mealtimes to worse glycemic control in individuals with diabetes (29). While these studies highlight the metabolic consequences of irregular or delayed eating, our study provides a novel contribution by directly quantifying daily eating irregularity and its 24-hour glycemic effects. These findings emphasize the importance of consistent meal timing as a strategy for improving glycemic control.

Carbohydrate intake plays a critical role in glycemic control among individuals with type 2 diabetes, with our novel evidence highlighting the time-dependent effects of carbohydrates on glucose dynamics. This is consistent with the previous study using CGM devices, where low-carbohydrate diets have been shown to increase glycemic TIR and CV in populations with type 2 diabetes (30). Significant associations between carbohydrate intake and glucose levels were observed across both in-range and variability CGM metrics in our study. Notably, our findings revealed that the effects of carbohydrate consumption were most pronounced around 10:00, likely reflecting postprandial glucose excursions following breakfast. This aligns with prior evidence showing that reducing the carbohydrate content in breakfast improves glucose control more effectively than adjusting lunch or dinner (31), and that a low-carbohydrate breakfast reduces 24-hour glycemic AUC and MAGE (32). Collectively, these findings emphasize the importance of targeting carbohydrate intake, particularly during breakfast, to optimize glycemic outcomes.

No significant associations were observed between dietary protein, lipid, or fiber intake and CGM metrics, yet FDA analyses revealed temporal effects. Protein intake showed a weak positive association with nocturnal glucose levels, potentially reflecting altered protein metabolism during extended fasting periods, as proposed by one previous study (33). FDA detected minor positive associations between total lipid intake and glucose levels in the afternoon. This observation may be influenced by the varying effects of fat types on glucose metabolism, as suggested by prior research (34). Our analysis examined total lipid intake as a whole to provide a broader perspective on overall dietary patterns and their relationship with glucose levels, rather than focusing on specific differences in fat quality. For dietary fiber, FDA revealed a significant negative association with glucose levels around noon. Although stronger associations have been reported previously (35), the discrepancy may reflect differences in diabetes cohorts (type 1 diabetes versus type 2 diabetes) or dietary patterns (Western versus Chinese diets). These findings highlight the utility of FDA in capturing time-specific associations between macronutrient intake and glycemic dynamics that may be missed by summary-level analyses.

Dietary vitamin D intake was identified as a significant factor influencing glycemic outcomes, particularly glycemic AUC. FDA analyses further showed pronounced effects of dietary vitamin D intake around noon and during the night. Our findings align with prior research showing that vitamin D-rich foods can reduce blood glucose levels in individuals with type 2 diabetes (36). While most previous studies have focused on serum vitamin D concentrations, our findings provide direct evidence of the impact of dietary vitamin D on glycemic parameters. These findings offer actionable insights for nutritional interventions to improve glycemic control.

Advanced statistical approaches from FDA uncovered novel insights into the temporal effects of TRE and nutritional content on glucose levels. Conventional CGM-derived metrics may fail to account for the multilevel structure of CGM data (13), and may lead to inconsistent conclusions if the appropriate metrics are not selected (37). Our application of function-on-scalar regression overcame these limitations and offered a more detailed understanding of glycemic profiles. This approach aligns with previous studies that used FDA techniques to examine the relationship between CGM levels and large-for-gestational-age infants in women with gestational diabetes (38–40). However, to our knowledge, no prior studies have employed function-on-scalar regression models to investigate the temporal effects of TRE on glucose levels. The use of FDA to analyze associations between nutritional intake and CGM data is also limited, with the only prior study focusing on nutrient-CGM associations during a 6-hour post-dinner period (13). Our study extends this analysis to a comprehensive 24-hour analysis of nutrient-glucose relationships, revealing distinct temporal patterns for different nutrients. While previous research has described postprandial responses to carbohydrates, lipids, and fiber, our analysis identifies precise temporal windows of nutrient effects and includes an investigation of dietary vitamin D, a nutrient found to be inadequate among the Chinese population (41).

This study is the first to examine the interplay between carbohydrate intake and a 10-hour eating window (TRE10) in individuals with type 2 diabetes. We found that while TRE10 slightly attenuated carbohydrate-induced glucose spikes in the morning, it amplified the glucose response in the afternoon. The observed afternoon amplification may reflect a misalignment between food intake timing and the body’s endogenous circadian regulation of glucose metabolism. This differential response pattern aligns with diurnal variations in glucose metabolism, where glucose tolerance is significantly impaired in the evening compared to the morning (42). Supporting this, a recent systematic review reported that early TRE is more effective than late TRE in improving metabolic health among overweight and obese individuals (43). Collectively, these findings underscore the importance of chrono-nutrition in diabetes management and suggest that aligning carbohydrate intake with circadian rhythms may enhance metabolic outcomes.

This study has several strengths, including the introduction of novel digital biomarkers (TRE10 and TWD) to quantify eating patterns and their irregularity, and the application of advanced statistical models from FDA to explore temporal glucose dynamics beyond conventional CGM summary metrics. By integrating CGM data with dietary intake, we identified time-dependent effects of TRE and nutrient consumption on glycemic control. These findings contribute to the growing field of chrono-nutrition and support the development of personalized diabetes management strategies. However, this study is not without limitations. First, the observational design precludes causal inference, meaning that the identified associations between TRE10, TWD, and glycemic outcomes cannot be interpreted as evidence of direct effects. Further research into the causal relationship between short eating windows and glycemic control among individuals with type 2 diabetes should be conducted. Second, the reliance on participants’ habitual eating patterns restricted our ability to explore alternative TRE cut-off hours or to differentiate between early and late TRE, both of which may have differing metabolic impacts. Controlled intervention studies are needed to evaluate these variations and identify the optimal timing and duration of eating windows. Additionally, our Chinese cohort may have unique culture, and future work should compare populations with different dietary habits, eating patterns, and cultural contexts.

## Acknowledgements

All authors report no conflicts of interest.

## Data Availability

Data access to the ShanghaiT2DM Dataset are available from the repository at https://figshare.com/collections/Diabetes_Datasets_ShanghaiT1DM_and_ShanghaiT2DM/6310860/2

## Funding

This study was supported by City University of Hong Kong internal research grant #9610473, the Institute of Digital Medicine and the Hong Kong Innovation and Technology Commission (InnoHK Project CIMDA). The funders played no role in the design, analysis, or interpretation of the results in this study.

## Notes

### Competing Interest Statement

The authors have declared no competing interest.

### Summary of Updates

Title revised for clarification; supplemental files updated.

