## Supplementary Figure, Supplementary Table for "Time-Restricted Eating and Dietary Intake Shape 24-Hour Glycemic Patterns in Type 2 Diabetes: A Functional Data Analysis Approach"

**Supplementary Material**

**Supplementary Figure 1.** Flowchart of participants from ShanghaiT2DM

**Supplementary Table 1.** Independent associations of time-restricted eating and nutritional contents with CGM-derived summary statistics

**Supplementary Table 2.** Subgroup analysis of independent associations between time-restricted eating and nutritional contents with CGM summary statistics among males

**Supplementary Table 3.** Subgroup analysis of independent associations between time-restricted eating and nutritional contents with CGM summary statistics among females

**Supplementary Table 4.** Subgroup analysis of independent associations between time-restricted eating and nutritional contents with CGM summary statistics among participants aged 63 (median) or above

**Supplementary Table 5.** Subgroup analysis of independent associations between time-restricted eating and nutritional contents with CGM summary statistics among participants aged below 63 (median)

**Supplementary Figure 1.** Flowchart of participants from ShanghaiT2DM

Exclusion:

- Insufficient wearing time of continuous glucose monitor (n = 4)
- Missing covariates or dietary records (n = 6)

Participants in final analysis

N = 90

ShanghaiT2DM Participants

N = 100

**Supplementary Table 1.** Independent associations of time-restricted eating and nutritional contents with CGM-derived summary statistics

| CGM-derived Summary Statistics | Exposures | $\boldsymbol{\beta(95\% CI)}$ | P value |
| --- | --- | --- | --- |
| TIR | TRE10 | 1.74 (-1.13, 4.6) | 0.235 |
|  | TWD (hours) | -0.61 (-1.45, 0.22) | 0.148 |
|  | Carbohydrate (10g) | -0.31 (-0.58, -0.03) | 0.029 |
|  | Protein (10g) | 0.24 (-0.19, 0.67) | 0.270 |
|  | Lipid (10g) | 0.08 (-0.57, 0.73) | 0.809 |
|  | Fiber (g) | -0.11 (-0.34, 0.13) | 0.373 |
|  | Vitamin D ($\mu$g) | 0.13 (-0.08, 0.33) | 0.221 |
| TAR | TRE10 | -2.11 (-4.88, 0.67) | 0.137 |
|  | TWD (hours) | 0.43 (-0.38, 1.24) | 0.294 |
|  | Carbohydrate (10g) | 0.28 (0.01, 0.55) | 0.039 |
|  | Protein (10g) | -0.25 (-0.67, 0.17) | 0.242 |
|  | Lipid (10g) | -0.09 (-0.71, 0.54) | 0.789 |
|  | Fiber (g) | 0.07 (-0.16, 0.30) | 0.528 |
|  | Vitamin D ($\mu$g) | -0.17 (-0.37, 0.03) | 0.094 |
| TBR | TRE10 | 0.40 (-0.52, 1.32) | 0.394 |
|  | TWD (hours) | 0.18 (-0.09, 0.45) | 0.183 |
|  | Carbohydrate (10g) | 0.03 (-0.06, 0.12) | 0.533 |
|  | Protein (10g) | 0.01 (-0.13, 0.15) | 0.903 |
|  | Lipid (10g) | 0.01 (-0.20, 0.22) | 0.942 |
|  | Fiber (g) | 0.03 (-0.05, 0.11) | 0.429 |
|  | Vitamin D ($\mu$g) | 0.04 (-0.02, 0.11) | 0.185 |
| AUC | TRE10 | -2.38 (-6.27, 1.51) | 0.230 |
|  | TWD (hours) | 0.22 (-0.90, 1.35) | 0.696 |
|  | Carbohydrate (10g) | 0.32 (-0.06, 0.69) | 0.103 |
|  | Protein (10g) | -0.13 (-0.72, 0.46) | 0.667 |
|  | Lipid (10g) | 0.49 (-0.39, 1.37) | 0.274 |
|  | Fiber (g) | 0.08 (-0.24, 0.40) | 0.614 |
|  | Vitamin D ($\mu$g) | -0.29 (-0.56, -0.01) | 0.040 |
| CV | TRE10 | -1.03 (-2.25, 0.20) | 0.100 |
|  | TWD (hours) | 0.36 (0, 0.72) | 0.050 |
|  | Carbohydrate (10g) | 0.16 (0.05, 0.28) | 0.005 |
|  | Protein (10g) | -0.10 (-0.28, 0.08) | 0.282 |
|  | Lipid (10g) | -0.14 (-0.41, 0.14) | 0.324 |
|  | Fiber (g) | 0.06 (-0.04, 0.16) | 0.223 |
|  | Vitamin D ($\mu$g) | 0 (-0.09, 0.09) | 0.970 |
| SD | TRE10 | -2.16 (-4.16, -0.15) | 0.035 |
|  | TWD (hours) | 0.44 (-0.15, 1.03) | 0.145 |
|  | Carbohydrate (10g) | 0.27 (0.08, 0.46) | 0.005 |
|  | Protein (10g) | -0.12 (-0.42, 0.18) | 0.420 |
|  | Lipid (10g) | -0.04 (-0.48, 0.41) | 0.878 |
|  | Fiber (g) | 0.07 (-0.09, 0.24) | 0.382 |
|  | Vitamin D ($\mu$g) | -0.05 (-0.20, 0.09) | 0.459 |
| MAGE | TRE10 | -7.14 (-13.62, -0.67) | 0.031 |
|  | TWD (hours) | 0.66 (-1.27, 2.59) | 0.501 |
|  | Carbohydrate (10g) | 0.71 (0.12, 1.31) | 0.018 |
|  | Protein (10g) | -0.23 (-1.20, 0.74) | 0.642 |
|  | Lipid (10g) | -0.01 (-1.44, 1.43) | 0.994 |
|  | Fiber (g) | 0.22 (-0.31, 0.75) | 0.408 |
|  | Vitamin D ($\mu$g) | -0.20 (-0.67, 0.27) | 0.394 |

The results of linear mixed models after adjusting age, gender, diabetes duration, BMI, alcohol, and smoking. TRE10 defined eating windows into "short" ($<$10 hours) and "long" ($\geq$10 hours). TWD defined as the time difference between the observed daily eating window and the participant's median eating window. All nutritional variables are daily dietary intake. AUC, hourly average area under the glucose curve; CV, coefficient of variation; MAGE, mean amplitude of glycemic excursions; SD, standard deviation; TAR, time above range (180 mg/dL); TBR, time below range (70 mg/dL); TIR, time in range (70 – 180 mg/dL).

**Supplementary Table 2.** Subgroup analysis of independent associations between time-restricted eating and nutritional contents with CGM summary statistics among males

| CGM-derived Summary Statistics | Exposures | $\boldsymbol{\beta(95\% CI)}$ | P value |
| --- | --- | --- | --- |
| TIR | TRE10 | 0.46 (-3.56, 4.49) | 0.821 |
|  | TWD (hours) | -1.07 (-2.45, 0.31) | 0.126 |
|  | Carbohydrate (10g) | -0.50 (-0.89, -0.12) | 0.011 |
|  | Protein (10g) | 0.89 (0.26, 1.52) | 0.006 |
|  | Lipid (10g) | 1.10 (0.16, 2.05) | 0.022 |
|  | Fiber (g) | -0.10 (-0.43, 0.23) | 0.559 |
|  | Vitamin D ($\mu$g) | 0.25 (-0.03, 0.54) | 0.082 |
| TAR | TRE10 | -0.68 (-4.62, 3.26) | 0.736 |
|  | TWD (hours) | 1.04 (-0.31, 2.39) | 0.130 |
|  | Carbohydrate (10g) | 0.42 (0.05, 0.80) | 0.028 |
|  | Protein (10g) | -0.95 (-1.56, -0.33) | 0.003 |
|  | Lipid (10g) | -1.13 (-2.06, -0.21) | 0.016 |
|  | Fiber (g) | 0.03 (-0.29, 0.36) | 0.848 |
|  | Vitamin D ($\mu$g) | -0.27 (-0.55, 0.01) | 0.055 |
| TBR | TRE10 | 0.27 (-0.72, 1.25) | 0.591 |
|  | TWD (hours) | 0.04 (-0.30, 0.38) | 0.813 |
|  | Carbohydrate (10g) | 0.09 (0, 0.19) | 0.061 |
|  | Protein (10g) | 0.07 (-0.09, 0.23) | 0.371 |
|  | Lipid (10g) | 0.06 (-0.18, 0.29) | 0.633 |
|  | Fiber (g) | 0.06 (-0.02, 0.14) | 0.156 |
|  | Vitamin D ($\mu$g) | 0.03 (-0.04, 0.10) | 0.465 |
| AUC | TRE10 | 0.09 (-5.15, 5.32) | 0.974 |
|  | TWD (hours) | 1.16 (-0.63, 2.95) | 0.202 |
|  | Carbohydrate (10g) | 0.51 (0.01, 1.02) | 0.047 |
|  | Protein (10g) | -1.01 (-1.83, -0.19) | 0.016 |
|  | Lipid (10g) | -0.96 (-2.19, 0.28) | 0.128 |
|  | Fiber (g) | 0.10 (-0.34, 0.53) | 0.655 |
|  | Vitamin D ($\mu$g) | -0.38 (-0.75, -0.01) | 0.045 |
| CV | TRE10 | -1.30 (-3.01, 0.42) | 0.137 |
|  | TWD (hours) | 0.45 (-0.15, 1.05) | 0.138 |
|  | Carbohydrate (10g) | 0.11 (-0.05, 0.27) | 0.173 |
|  | Protein (10g) | -0.14 (-0.41, 0.13) | 0.292 |
|  | Lipid (10g) | -0.34 (-0.73, 0.06) | 0.098 |
|  | Fiber (g) | 0 (-0.14, 0.14) | 0.968 |
|  | Vitamin D ($\mu$g) | 0.02 (-0.1, 0.14) | 0.732 |
| SD | TRE10 | -1.90 (-4.62, 0.81) | 0.169 |
|  | TWD (hours) | 0.83 (-0.11, 1.77) | 0.083 |
|  | Carbohydrate (10g) | 0.21 (-0.04, 0.47) | 0.104 |
|  | Protein (10g) | -0.39 (-0.82, 0.03) | 0.071 |
|  | Lipid (10g) | -0.65 (-1.28, -0.02) | 0.045 |
|  | Fiber (g) | -0.03 (-0.26, 0.19) | 0.776 |
|  | Vitamin D ($\mu$g) | -0.03 (-0.22, 0.17) | 0.778 |
| MAGE | TRE10 | -9.25 (-17.98, -0.51) | 0.038 |
|  | TWD (hours) | 1.92 (-1.16, 4.99) | 0.220 |
|  | Carbohydrate (10g) | 0.46 (-0.34, 1.26) | 0.256 |
|  | Protein (10g) | -0.85 (-2.22, 0.52) | 0.223 |
|  | Lipid (10g) | -1.73 (-3.75, 0.28) | 0.092 |
|  | Fiber (g) | -0.22 (-0.93, 0.50) | 0.557 |
|  | Vitamin D ($\mu$g) | 0.01 (-0.62, 0.64) | 0.983 |

The results of linear mixed models after adjusting age, diabetes duration, BMI, alcohol, and smoking. TRE10 defined eating windows into "short" ($<$10 hours) and "long" ($\geq$10 hours), with the "long" eating window group serving as the reference. TWD defined as the time difference between the observed daily eating window and the participant's median eating window. All nutritional variables are daily dietary intake. AUC, hourly average area under the glucose curve; CV, coefficient of variation; MAGE, mean amplitude of glycemic excursions; SD, standard deviation; TAR, time above range (180 mg/dL); TBR, time below range (70 mg/dL); TIR, time in range (70 – 180 mg/dL).

**Supplementary Table 3.** Subgroup analysis of independent associations between time-restricted eating and nutritional contents with CGM summary statistics among females

| CGM-derived Summary Statistics | Exposures | $\boldsymbol{\beta(95\% CI)}$ | P value |
| --- | --- | --- | --- |
| TIR | TRE10 | 3.22 (-0.84, 7.28) | 0.120 |
|  | TWD (hours) | -0.29 (-1.32, 0.74) | 0.580 |
|  | Carbohydrate (10g) | -0.08 (-0.48, 0.31) | 0.677 |
|  | Protein (10g) | -0.35 (-0.93, 0.23) | 0.237 |
|  | Lipid (10g) | -0.89 (-1.75, -0.02) | 0.045 |
|  | Fiber (g) | -0.14 (-0.47, 0.19) | 0.411 |
|  | Vitamin D ($\mu$g) | -0.02 (-0.31, 0.28) | 0.917 |
| TAR | TRE10 | -3.76 (-7.65, 0.13) | 0.058 |
|  | TWD (hours) | 0.01 (-0.98, 1.00) | 0.986 |
|  | Carbohydrate (10g) | 0.11 (-0.27, 0.49) | 0.576 |
|  | Protein (10g) | 0.4 (-0.16, 0.95) | 0.165 |
|  | Lipid (10g) | 0.9 (0.06, 1.73) | 0.035 |
|  | Fiber (g) | 0.14 (-0.18, 0.46) | 0.386 |
|  | Vitamin D ($\mu$g) | -0.05 (-0.33, 0.23) | 0.702 |
| TBR | TRE10 | 0.40 (-1.09, 1.89) | 0.598 |
|  | TWD (hours) | 0.36 (-0.04, 0.76) | 0.076 |
|  | Carbohydrate (10g) | -0.02 (-0.15, 0.12) | 0.776 |
|  | Protein (10g) | -0.05 (-0.26, 0.15) | 0.609 |
|  | Lipid (10g) | -0.02 (-0.33, 0.29) | 0.902 |
|  | Fiber (g) | -0.01 (-0.13, 0.11) | 0.817 |
|  | Vitamin D ($\mu$g) | 0.07 (-0.04, 0.19) | 0.190 |
| AUC | TRE10 | -5.26 (-11.07, 0.55) | 0.076 |
|  | TWD (hours) | -0.42 (-1.89, 1.04) | 0.570 |
|  | Carbohydrate (10g) | 0.09 (-0.48, 0.66) | 0.758 |
|  | Protein (10g) | 0.69 (-0.14, 1.53) | 0.102 |
|  | Lipid (10g) | 1.86 (0.62, 3.10) | 0.003 |
|  | Fiber (g) | 0.10 (-0.38, 0.58) | 0.690 |
|  | Vitamin D ($\mu$g) | -0.20 (-0.62, 0.22) | 0.344 |
| CV | TRE10 | -0.71 (-2.46, 1.05) | 0.430 |
|  | TWD (hours) | 0.29 (-0.16, 0.74) | 0.206 |
|  | Carbohydrate (10g) | 0.22 (0.05, 0.38) | 0.010 |
|  | Protein (10g) | -0.06 (-0.31, 0.19) | 0.617 |
|  | Lipid (10g) | 0.04 (-0.33, 0.42) | 0.819 |
|  | Fiber (g) | 0.14 (-0.01, 0.28) | 0.061 |
|  | Vitamin D ($\mu$g) | -0.02 (-0.15, 0.11) | 0.747 |
| SD | TRE10 | -2.14 (-5.09, 0.81) | 0.154 |
|  | TWD (hours) | 0.14 (-0.63, 0.91) | 0.719 |
|  | Carbohydrate (10g) | 0.36 (0.09, 0.63) | 0.010 |
|  | Protein (10g) | 0.15 (-0.27, 0.56) | 0.494 |
|  | Lipid (10g) | 0.57 (-0.05, 1.19) | 0.074 |
|  | Fiber (g) | 0.22 (-0.02, 0.46) | 0.072 |
|  | Vitamin D ($\mu$g) | -0.09 (-0.30, 0.13) | 0.436 |
| MAGE | TRE10 | -4.32 (-13.91, 5.27) | 0.376 |
|  | TWD (hours) | -0.38 (-2.89, 2.13) | 0.765 |
|  | Carbohydrate (10g) | 0.97 (0.09, 1.84) | 0.032 |
|  | Protein (10g) | 0.35 (-1.00, 1.71) | 0.609 |
|  | Lipid (10g) | 1.62 (-0.39, 3.62) | 0.113 |
|  | Fiber (g) | 0.74 (-0.03, 1.51) | 0.059 |
|  | Vitamin D ($\mu$g) | -0.47 (-1.17, 0.24) | 0.196 |

Since all females in the study are non-drinkers and non-smokers, the linear mixed models were adjusted for age, diabetes duration, and BMI. TRE10 defined eating windows into "short" ($<$10 hours) and "long" ($\geq$10 hours), with the "long" eating window group serving as the reference. TWD defined as the time difference between the observed daily eating window and the participant's median eating window. All nutritional variables are daily dietary intake. AUC, hourly average area under the glucose curve; CV, coefficient of variation; MAGE, mean amplitude of glycemic excursions; SD, standard deviation; TAR, time above range (180 mg/dL); TBR, time below range (70 mg/dL); TIR, time in range (70 – 180 mg/dL).

**Supplementary Table 4.** Subgroup analysis of independent associations between time-restricted eating and nutritional contents with CGM summary statistics among participants aged 63 (median) or above

| CGM-derived Summary Statistics | Exposures | $\boldsymbol{\beta(95\% CI)}$ | P value |
| --- | --- | --- | --- |
| TIR | TRE10 | 0.63 (-3.53, 4.79) | 0.766 |
|  | TWD (hours) | -0.21 (-1.39, 0.97) | 0.725 |
|  | Carbohydrate (10g) | -0.13 (-0.53, 0.26) | 0.514 |
|  | Protein (10g) | 0.03 (-0.58, 0.65) | 0.918 |
|  | Lipid (10g) | 0.02 (-0.87, 0.90) | 0.972 |
|  | Fiber (g) | -0.07 (-0.39, 0.25) | 0.660 |
|  | Vitamin D ($\mu$g) | 0.14 (-0.13, 0.40) | 0.312 |
| TAR | TRE10 | -0.95 (-5.08, 3.18) | 0.652 |
|  | TWD (hours) | -0.04 (-1.21, 1.12) | 0.940 |
|  | Carbohydrate (10g) | 0.05 (-0.34, 0.45) | 0.793 |
|  | Protein (10g) | -0.07 (-0.68, 0.54) | 0.827 |
|  | Lipid (10g) | -0.01 (-0.89, 0.88) | 0.989 |
|  | Fiber (g) | 0.08 (-0.23, 0.40) | 0.606 |
|  | Vitamin D ($\mu$g) | -0.18 (-0.44, 0.08) | 0.186 |
| TBR | TRE10 | 0.16 (-0.94, 1.26) | 0.772 |
|  | TWD (hours) | 0.26 (-0.08, 0.60) | 0.132 |
|  | Carbohydrate (10g) | 0.10 (0, 0.20) | 0.052 |
|  | Protein (10g) | 0.03 (-0.14, 0.19) | 0.731 |
|  | Lipid (10g) | 0 (-0.23, 0.23) | 0.999 |
|  | Fiber (g) | 0.02 (-0.07, 0.1) | 0.696 |
|  | Vitamin D ($\mu$g) | 0.04 (-0.04, 0.11) | 0.332 |
| AUC | TRE10 | -1.40 (-7.28, 4.47) | 0.639 |
|  | TWD (hours) | -0.57 (-2.23, 1.09) | 0.501 |
|  | Carbohydrate (10g) | -0.14 (-0.70, 0.42) | 0.632 |
|  | Protein (10g) | 0.25 (-0.62, 1.12) | 0.575 |
|  | Lipid (10g) | 0.45 (-0.8, 1.71) | 0.480 |
|  | Fiber (g) | 0 (-0.45, 0.45) | 0.992 |
|  | Vitamin D ($\mu$g) | -0.27 (-0.64, 0.1) | 0.158 |
| CV | TRE10 | -0.45 (-2.25, 1.35) | 0.624 |
|  | TWD (hours) | 0.29 (-0.24, 0.82) | 0.282 |
|  | Carbohydrate (10g) | 0.19 (0.02, 0.35) | 0.026 |
|  | Protein (10g) | -0.07 (-0.33, 0.20) | 0.618 |
|  | Lipid (10g) | -0.06 (-0.44, 0.32) | 0.745 |
|  | Fiber (g) | -0.01 (-0.15, 0.13) | 0.848 |
|  | Vitamin D ($\mu$g) | 0.02 (-0.10, 0.14) | 0.750 |
| SD | TRE10 | -1.37 (-4.57, 1.83) | 0.401 |
|  | TWD (hours) | 0.17 (-0.76, 1.10) | 0.720 |
|  | Carbohydrate (10g) | 0.19 (-0.11, 0.48) | 0.219 |
|  | Protein (10g) | -0.03 (-0.50, 0.45) | 0.910 |
|  | Lipid (10g) | 0.04 (-0.64, 0.72) | 0.905 |
|  | Fiber (g) | -0.04 (-0.29, 0.21) | 0.752 |
|  | Vitamin D ($\mu$g) | -0.03 (-0.23, 0.18) | 0.802 |
| MAGE | TRE10 | -4.84 (-14.46, 4.78) | 0.323 |
|  | TWD (hours) | 0.26 (-2.67, 3.18) | 0.863 |
|  | Carbohydrate (10g) | 0.64 (-0.24, 1.51) | 0.152 |
|  | Protein (10g) | -0.15 (-1.58, 1.29) | 0.839 |
|  | Lipid (10g) | 0.08 (-1.95, 2.11) | 0.941 |
|  | Fiber (g) | 0.06 (-0.69, 0.81) | 0.881 |
|  | Vitamin D ($\mu$g) | -0.23 (-0.88, 0.41) | 0.474 |

The results of linear mixed models after adjusting gender, diabetes duration, BMI, alcohol, and smoking. TRE10 defined eating windows into "short" ($<$10 hours) and "long" ($\geq$10 hours), with the "long" eating window group serving as the reference. TWD defined as the time difference between the observed daily eating window and the participant's median eating window. All nutritional variables are daily dietary intake. T AUC, hourly average area under the glucose curve; CV, coefficient of variation; MAGE, mean amplitude of glycemic excursions; SD, standard deviation; TAR, time above range (180 mg/dL); TBR, time below range (70 mg/dL); TIR, time in range (70 – 180 mg/dL). IR, time in range (70 – 180 mg/dL); TAR, time above range (180 mg/dL); TBR, time below range (70 mg/dL); AUC, hourly average area under the glucose curve; CV, coefficient of variation; MAGE, mean amplitude of glycaemic excursions; SD, standard deviation.

**Supplementary Table 5.** Subgroup analysis of independent associations between time-restricted eating and nutritional contents with CGM summary statistics among participants aged below 63 (median)

| CGM-derived Summary Statistics | Exposures | $\boldsymbol{\beta(95\% CI)}$ | P value |
| --- | --- | --- | --- |
| TIR | TRE10 | 2.68 (-1.32, 6.67) | 0.189 |
|  | TWD (hours) | -0.96 (-2.15, 0.22) | 0.111 |
|  | Carbohydrate (10g) | -0.46 (-0.85, -0.07) | 0.020 |
|  | Protein (10g) | 0.44 (-0.17, 1.05) | 0.159 |
|  | Lipid (10g) | 0.16 (-0.78, 1.11) | 0.733 |
|  | Fiber (g) | -0.15 (-0.50, 0.20) | 0.402 |
|  | Vitamin D ($\mu$g) | 0.12 (-0.20, 0.44) | 0.453 |
| TAR | TRE10 | -3.03 (-6.81, 0.76) | 0.116 |
|  | TWD (hours) | 0.86 (-0.26, 1.98) | 0.134 |
|  | Carbohydrate (10g) | 0.48 (0.11, 0.85) | 0.010 |
|  | Protein (10g) | -0.42 (-1.00, 0.16) | 0.156 |
|  | Lipid (10g) | -0.19 (-1.09, 0.71) | 0.683 |
|  | Fiber (g) | 0.07 (-0.27, 0.40) | 0.696 |
|  | Vitamin D ($\mu$g) | -0.17 (-0.47, 0.14) | 0.286 |
| TBR | TRE10 | 0.44 (-0.94, 1.81) | 0.532 |
|  | TWD (hours) | 0.10 (-0.30, 0.51) | 0.617 |
|  | Carbohydrate (10g) | -0.01 (-0.15, 0.13) | 0.868 |
|  | Protein (10g) | -0.01 (-0.22, 0.20) | 0.927 |
|  | Lipid (10g) | 0.04 (-0.29, 0.37) | 0.801 |
|  | Fiber (g) | 0.08 (-0.05, 0.20) | 0.221 |
|  | Vitamin D ($\mu$g) | 0.05 (-0.06, 0.16) | 0.341 |
| AUC | TRE10 | -3.08 (-8.28, 2.12) | 0.244 |
|  | TWD (hours) | 0.96 (-0.58, 2.50) | 0.222 |
|  | Carbohydrate (10g) | 0.70 (0.19, 1.21) | 0.008 |
|  | Protein (10g) | -0.48 (-1.28, 0.32) | 0.239 |
|  | Lipid (10g) | 0.48 (-0.76, 1.72) | 0.447 |
|  | Fiber (g) | 0.17 (-0.29, 0.62) | 0.477 |
|  | Vitamin D ($\mu$g) | -0.32 (-0.73, 0.10) | 0.136 |
| CV | TRE10 | -1.47 (-3.13, 0.18) | 0.080 |
|  | TWD (hours) | 0.39 (-0.10, 0.88) | 0.121 |
|  | Carbohydrate (10g) | 0.13 (-0.02, 0.29) | 0.097 |
|  | Protein (10g) | -0.14 (-0.39, 0.11) | 0.269 |
|  | Lipid (10g) | -0.22 (-0.61, 0.17) | 0.263 |
|  | Fiber (g) | 0.14 (0, 0.28) | 0.056 |
|  | Vitamin D ($\mu$g) | -0.02 (-0.15, 0.12) | 0.795 |
| SD | TRE10 | -2.71 (-5.15, -0.26) | 0.030 |
|  | TWD (hours) | 0.64 (-0.08, 1.37) | 0.082 |
|  | Carbohydrate (10g) | 0.32 (0.09, 0.55) | 0.008 |
|  | Protein (10g) | -0.24 (-0.61, 0.14) | 0.211 |
|  | Lipid (10g) | -0.17 (-0.74, 0.41) | 0.573 |
|  | Fiber (g) | 0.19 (-0.02, 0.4) | 0.083 |
|  | Vitamin D ($\mu$g) | -0.09 (-0.28, 0.11) | 0.384 |
| MAGE | TRE10 | -8.64 (-17.12, -0.15) | 0.046 |
|  | TWD (hours) | 0.83 (-1.70, 3.37) | 0.518 |
|  | Carbohydrate (10g) | 0.74 (-0.05, 1.53) | 0.067 |
|  | Protein (10g) | -0.33 (-1.62, 0.95) | 0.608 |
|  | Lipid (10g) | -0.14 (-2.13, 1.84) | 0.886 |
|  | Fiber (g) | 0.42 (-0.31, 1.15) | 0.260 |
|  | Vitamin D ($\mu$g) | -0.13 (-0.82, 0.55) | 0.701 |

The results of linear mixed models after adjusting gender, diabetes duration, BMI, alcohol, and smoking. TRE10 defined eating windows into "short" ($<$10 hours) and "long" ($\geq$10 hours), with the "long" eating window group serving as the reference. TWD defined as the time difference between the observed daily eating window and the participant's median eating window. All nutritional variables are daily dietary intake. AUC, hourly average area under the glucose curve; CV, coefficient of variation; MAGE, mean amplitude of glycemic excursions; SD, standard deviation; TAR, time above range (180 mg/dL); TBR, time below range (70 mg/dL); TIR, time in range (70 – 180 mg/dL).
